# An increase in C-reactive protein levels during antidepressant treatment predicts treatment non-response in major depressive disorder

**DOI:** 10.1101/2025.08.27.25331881

**Authors:** Marius Etzel, Yevgenia Rosenblum, Martin Dresler, Axel Steiger, Thorsten Mikoteit, Marcel Zeising

## Abstract

**Background:** One-third of patients with major depressive disorder (MDD) exhibit low-grade inflammation as reflected by C-reactive protein (CRP) concentrations > 3 mg/L. We explored whether CRP changes from baseline to week one of antidepressant treatment (ΔCRP) can serve as a marker of treatment response.

**Methods:** CRP plasma levels were measured at baseline and after the first week of treatment in 33 MDD patients and correlated with patients’ Hamilton depression scale (HAM-D), while adjusting for age, gender and body mass index. The early and final antidepressant responses were defined as a > 25% and > 50% HAM-D score reduction at week one and four of treatment compared to baseline, respectively. We compared baseline and week-one CRP levels with the paired t-test within responders and non-responders separately and ΔCRP between the groups with the ANCOVA.

**Results:** Higher ΔCRP correlated with lower final ΔHAM-D scores (r = −0.5, p = 0.006). Non-responders showed higher ΔCRP – but not baseline and week one CRP – than responders (p = 0.018, Cohen’s d = 1.1). A ΔCRP increase was observed in 13/16 (81%) non-responders and 7/17 (41%) responders (Fisher’s exact test’s p = 0.03). A ΔCRP increase combined with an early non-response was observed in 13/16 (81%) non-responders and 1/17 (6%) responders (p < 0.0001).

**Conclusions:** Rather early ΔCRP at week one than baseline CRP might be indicative of treatment response at week four, especially if combined with early ΔHAM-D. In the future, ΔCRP could be introduced into psychiatric practice to guide treatment plans.

## Introduction

Only around half of patients with major depressive disorder (MDD) respond to an initial four-week course of antidepressant treatment as evaluated by clinicians (Zajecka, 2003; Trivedi et al., 2006). The likelihood of complete remission is even lower, 30 – 40% (Iosifescu, 2022). Non-responders should enter another four-week period when they take a different antidepressant followed by an additional therapeutic assessment by a clinician. Such a trial-and-error approach results in delayed treatment improvement and a higher disease burden.

It is widely agreed that a reliable biological marker is needed to guide clinicians in antidepressant prescriptions. Previous studies tested clinical, electroencephalographic, neuroimaging, pharmacogenomic and inflammatory markers, yet none of them has been introduced in daily psychiatric practice (Strawbridge et al., 2018; Iosifescu, 2022). Here, we explored whether plasma levels of the C-reactive protein (CRP) can serve as a marker of the antidepressant treatment response.

CRP is a nonspecific acute-phase protein synthesized in the liver in response to stimulation from the proinflammatory cytokines interleukin-6 and interleukin-1 (Young et al., 2014). CRP is widely used in clinical practice as a marker of inflammation. It is accessible by a commonly available, inexpensive blood test that gives reproducible results and is quite stable across 24 hours (Chamberlain et al., 2019). There is evidence that MDD patients – and even more so patients with treatment-resistant depression – have elevated CRP levels compared to controls (Howren et al., 2009; Strawbridge et al., 2018; Chamberlain et al., 2019).

Given that inflammatory processes are involved in the onset, maintenance and remission of depressive symptoms, several studies explored whether inflammation markers could guide MDD diagnosis and therapy (Young et al., 2014). Moreover, recently, the immunological model of MDD has been introduced to explain some of the MDD symptoms as the non-specific reaction of the organism to infection and inflammation (Pastis et al., 2024). Likewise, the existence of the inflammatory subtype of MDD has been suggested (Miller et al., 2025). In view of the ability of proinflammatory cytokines to reduce the extracellular availability of serotonin, several studies further explored whether inflammation markers, including CRP, could be used as predictors of antidepressant treatment response.

Here, we measured within-subject changes in CRP levels from baseline to the end of the first week of antidepressant treatment (herein referred to as ΔCRP) and examined whether they could predict patients’ treatment response. We hypothesized that a within-subject decrease (or no change when there is no inflammation at baseline) in CRP levels after the onset of antidepressant treatment is an indicator of treatment response, while an increase (or no change when there is baseline inflammation) in CRP levels is a marker of treatment non-response. This hypothesis is based on reports that patients who responded to treatment showed lower proinflammatory/anti-inflammatory protein ratios compared to non-responders or healthy controls (Young et al., 2016). Currently, there is limited evidence on changes in CRP during the very early treatment phase and most prior studies measured the concentrations of the inflammation markers before starting medication treatment rather than ΔCRP with promising but inconclusive results (Strawbridge et al., 2018). Given that inflammation is often associated with different comorbidities, such as metabolic disorders, age, higher numbers of previous depressive episodes and the atypical type of depression (Young et al., 2016) together with high between-subject heterogeneity of MDD patients, we expect that the use of ΔCRP, i.e., the repeated within-subject measure design, will be more sensitive than the between-subject analysis that has been used in literature thus far with low sensitivity and specificity of the results. This, in turn, limited the use of inflammation markers to the research environment only and prevented their utilization in clinical settings. We expect that the use of ΔCRP will address this issue.

## Methods and Materials

### Participants

We recruited 63 consecutive in- and outpatients with MDD of both sexes from daily clinical practice at the Max Planck Institute of Psychiatry in Munich, Germany, between 2013 and 2016. All patients met DSM-IV criteria for a first or recurrent episode of MDD as diagnosed by two senior psychiatrists and were scored on the 21-item Hamilton depression rating scale (HAM-D) (Hamilton, 1960) ≥ 14. Likewise, we recruited 58 healthy volunteers with no current or past psychiatric disorders who were paid for participation in the study. The study was approved by the Ethics Committee of the Ludwig Maximilian University of Munich. All participants provided written informed consent.

All patients started medication treatment according to the psychiatrist’s choice within a few days after admission. Plasma concentrations of antidepressant medication were monitored weekly to ensure clinically efficient drug levels.

Exclusion criteria included pregnancy, history of drug/alcohol dependance, history of head trauma, serious risk of suicide, personality disorder and severe somatic diseases as well as long-acting medication including fluoxetine and depot neuroleptics, and if they underwent therapeutic sleep deprivation, electroconvulsive therapy, shift work or transmeridian travel within the three months prior to the study. Benzodiazepine or non-benzodiazepine anxiolytics were allowed in low stable dosages.

21 patients dropped out from the study because they either discontinued antidepressant therapy prematurely (n = 3), did not complete EEG or MRI assessments (n = 6), had restless leg syndrome after mirtazapine (n = 1), or because of non-compliance to the study (n = 11). 8 patients could not be included in the analyses because their CRP data were missing and one – because he was later diagnosed with schizophrenia. This resulted in the final sample size of 33 patients (age: 30.7 ± 9.9 years, 17 males).

### Study design

This report is a part of a larger unpublished study that had the following design (Fig.1). All participants underwent a physical examination, routine blood and urine diagnostics, preliminary electrocardiogram, electroencephalogram and cranial magnetic resonance imaging to screen for underlying and secondary conditions. Patients spent two consecutive night (23:00 to 07:00) in the sleep laboratory before the treatment onset, and two additional consecutive nights one week after the treatment. Patients filled the Pittsburgh Sleep Quality Index questionnaire and HAM-D in the morning upon awakening after the experimental night in the sleep lab. Then, blood sampling to assess CRP was taken.

**Figure 1.**
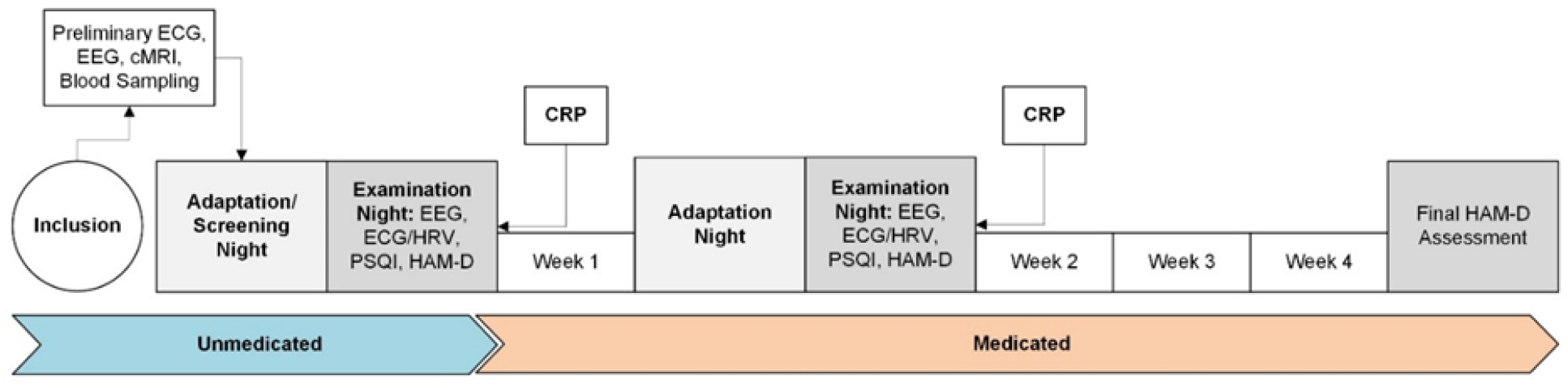
Study design. PSQI – Pittsburgh Sleep Quality Index, HAM-D – Hamilton depression rating scale, HRV - heart rate variability, ECG – electrocardiogram, CRP – C-reactive protein.

CRP was assessed two times: before starting medication and after the first week of treatment. HAM-D was assessed three times: before starting medication and after the first and fourth weeks of treatment.

### CRP

Venous blood sampling was performed at 8:30 in the morning. Patients were fasting and in a relaxed supine position. Serum-Gel-S-Monovettes® (Sarstedt AG & Co, Nümbrecht, Germany) were used. After collection, the blood samples were immediately processed by the in-house laboratory of the Max Planck Institute of Psychiatry, which adheres to the guidelines of the German Medical Association. Quality control is maintained by participating in interlaboratory ring trials five times a year.

CRP values were measured using a latex-enhanced immunological turbidity assay (cobas® CRPL3 pack on cobas® c501, Roche Diagnostics, Mannheim, Germany). The analytical measurement range of this assay was 0.3 – 350 mg/L, with an inter-assay coefficient of variation (CV) of 1.7 – 11.1% and an intra-assay CV of 1.2 – 3.6% (Roche Diagnostics, 2017). The assay used in this investigation is classified as a wide-range CRP assay, which provides sufficient resolution for CRP values below 5 mg/L (“low-grade inflammation”), while maintaining a higher upper range and lower cost compared to high sensitivity CRP assays (Li et al., 2019; Ziv-Baran et al., 2017). For comparisons of this specific assay to high sensitivity CRP assays from the same manufacturer, see Li et al. (2019) and Monneret et al. (2018). No patients with CRP levels above 10 mg/L were excluded *a priori*, which is in line with current recommendations (Mac Giollabhui et al., 2020; Moriarity et al., 2021).

Five patients (two non-responders and three responders) had CRP levels below 0.3 mg/L (i.e., below the analytical range of the assay). Given that there were no clear indications in the literature whether CRP levels < 0.3 mg/L, i.e. almost 0 mg/L, should be excluded from analyses, we included them in our analyses. An additional analysis where those five patients were excluded yielded similar results (the findings are not shown, yet they could be easily calculated with the Supplemental Excel File and SPSS Syntax File that will be shared upon the publication of this Manuscript).

### Statistical analysis

To determine if the CRP data came from normal distribution we performed the Shapiro-Wilk test. We obtained p-values < 0.05, which indicated that the data was not normally distributed. To account for this, we performed a square root transformation of the obtained CRP values. After this transformation, the data showed normal distribution (p > 0.05). For all analyses described below, we used the transformed CRP values.

To assess how CRP levels relate to the corresponding HAM-D scores as well as how ΔCRP from baseline to week one correlates with final ΔHAM-D from baseline to week four, we performed partial Pearson’s correlations with adjustments for the patients’ age, gender and body mass index (BMI) as from literature these covariates are known as potential confounders of CRP levels in MDD (Horn et al., 2018). Smoking was not included as a covariate, since there were only 3/28 smokers. The antidepressant class was not included as a covariate, since there were 5 classes for 28 participants only. The Benjamini-Hochberg’s adjustment was applied to control for multiple comparisons (3 tests) with a false discovery rate set at 0.05 and the α-level set in the 0.017 – 0.050 range.

Next (as we obtained significant results), we stratified all patients by their final ΔHAM-D scores into responders and non-responders groups. The early response was defined as a > 25% reduction after one week, the final response – as a > 50% reduction in HAM-D after four weeks of antidepressant treatment compared to baseline.

For within-group analyses, we compared the baseline and week-one levels of CRP levels in each group separately using the Student’s two-tailed paired t-test. For between-subject analyses, we performed the ANCOVA with the “ΔCRP” as a dependent variable, the “group” (responders vs non-responders) – as the between-subject predicting factor, and the “gender”, “age”, and “BMI” as covariates. Effect sizes were assessed with Cohen’s d and partial eta squared.

To assess the ability of ΔCRP to discriminate between responders and non-responders, we calculated the area under the receiver operating characteristic (ROC) curve (AUC). The sensitivity and specificity of the tests were evaluated using the median ΔCRP of all participants as a cutoff (ΔCRP = 0.02 mg/L). We also calculated the corresponding positive predictive value, also known as precision, i.e., the number of “true positives” (here, responders) divided by the sum of the true and false positives, the negative predictive value, i.e., the number of “true negatives” (here, non-responders) divided by the sum of the true and false negatives), and accuracy, i.e., the sum of the true positives and true negatives divided by n.

To determine whether there exists a statistically significant association between the early and late antidepressant response/non-response and ΔCRP increase/no increase we used a 2×2 contingency table and Fisher’s Exact Test.

Matlab (version R2021b, The MathWorks, Inc., Natick, MA) and SPSS (version 25; IBM Corp.) were used for all statistical analyses.

## Results

First, we performed partial correlations between square root transformed CRP values and HAM-D scores with an adjustment for patients’ age, gender and BMI (Fig.2). We found that the higher the increase in CRP levels (indicating some increase in inflammation) from baseline to week one of treatment, the lower the change in the patients’ final HAM-D scores (reflecting the smaller improvement in depression symptoms) after the correction for multiple comparisons (r = −0.49, p = 0.006). Baseline and week-one CRP levels did not correlate with the corresponding HAM-D scores. When the correction for the effects of age, gender and BMI was not performed, besides the ΔCRP – ΔHAM-D correlation, we also observed a positive correlation between the CRP levels and HAM-D scores at week one after treatment (r = 0.4, p = 0.03).

**Figure 2.**
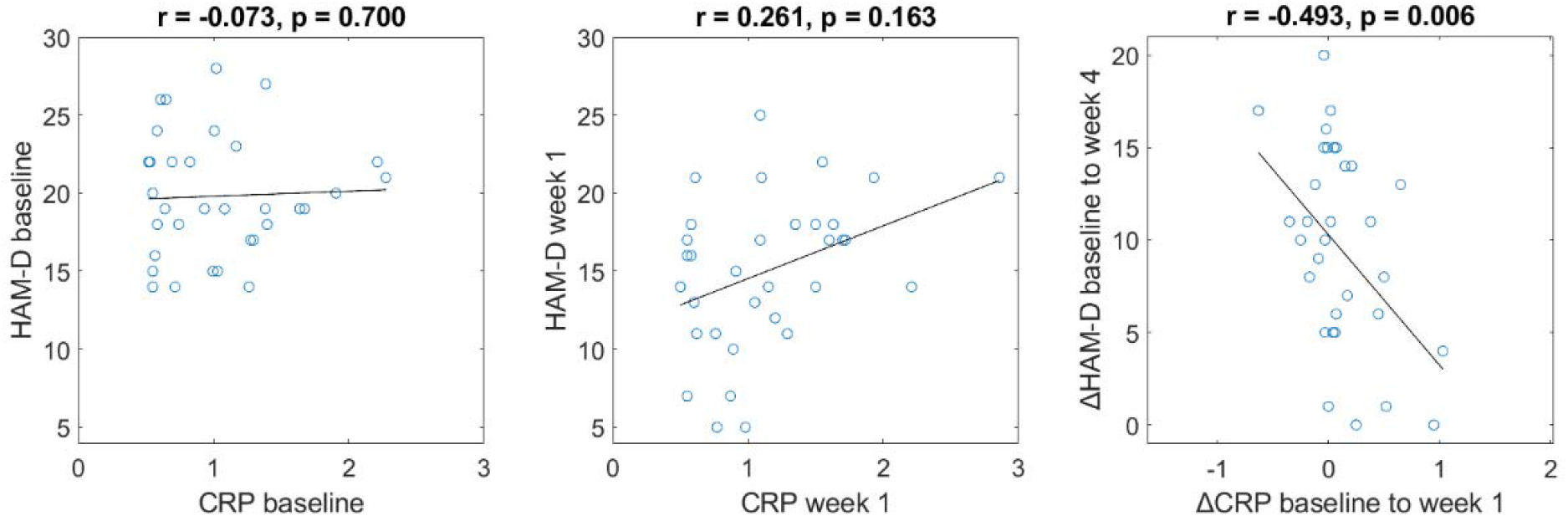
CRP vs HAM-D. The partial Pearson’s correlations adjusted for patients’ age, gender and BMI, where r’s are correlation coefficients. CRP levels at baseline (left) and week one (middle) did not correlate with the corresponding HAM-D scores. Right: a higher increase in ΔCRP from baseline to week one of treatment is correlated with smaller changes in patients’ final ΔHAM-D (i.e., no improvement in depression symptoms). Square root transformed CRP values are shown. Age range: 20 – 51 years, median: 28 years, n = 33, |r| < 0.3 are considered as weak correlations, 0.3 < |r| < 0.7 are considered as moderate scores.

Next, we divided all patients into responders and non-responders. Responders and non-responders had similar clinical and demographic characteristics at baseline (Table 1). We compared their CRP levels within and between the groups.

**Table 1.** Demographic and clinical variables of the participants at baseline.

| Characteristic | Non-responders<br>n = 16 | Responders<br>n = 17 |
| --- | --- | --- |
| Age, years | 31.1 ± 10.0 | 30.4 ± 10.0 |
| Gender: males, % | 50 | 53 |
| No. of previous depressive episodes | 1.3 ± 2.0 | 1.2 ± 1.3 |
| Age at 1<sup>st</sup> episode, years | 26.4 ± 8.6 | 25.4 ± 9.6 |
| Body Mass Index, kg/m<sup>2</sup> | 24.2 ± 4.1 | 24.8 ± 4.4 |
| Smokers, % | 12.5 | 6 |
| Antidepressants, %: |  |  |
| Selective serotonin-norepinephrine reuptake inhibitors (SNRI) | 6.3 | 11.8 |
| Selective serotonin reuptake inhibitors (SSRI) | 25 | 41 |
| Tricyclic antidepressants (TCA) | 25 | 17.6 |
| Noradrenergic and specific serotonergic antidepressant (NaSSA) | 12.5 | 23.5 |
| Norepinephrine-dopamine reuptake inhibitor (NDRI) | 31.3 | 5.9 |
| REM sleep suppressive antidepressants | 37.5 | 64.7 |

The between-group ANCOVA with adjustments for patients’ gender, age and BMI revealed a higher ΔCRP (i.e., a CRP increase from baseline) in non-responders compared to responders (F = 7.2, p = 0.012) with a large effect size (Cohen’s d = 1.16 or partial eta squared = 0.21). None of the covariates showed a significant effect on ΔCRP. Responders and non-responders had comparable CRP levels at baseline and week one (Table 2).

**Table 2.** CRP and HAM-D values of responders and non-responders (mean + std)

| Variable | Hamilton depression rating scale |  | C-reactive protein |  |
| --- | --- | --- | --- | --- |
| Time/Group | Non-responders<br>n = 16 | Responders<br>n = 17 | Non-responders<br>n = 16 | Responders<br>n = 17 |
| Baseline | 19.6 ± 4.3 | 20.0 ± 3.5 | 1.04 ± 0.84 | 1.56 ± 1.57 |
| Week one | 17.3 ± 3.2 | 12.9 ± 5.3 | 1.70 ± 1.26 | 1.54 ± 1.97 |
| Week four | 14.8 ± 3.9 | 6.1 ± 3.0 | --- | --- |
| Δ from baseline to week one | 2.3 ± 3.1 | 7.1 ± 4.7 | 0.65 ± 0.98 | -0.03 ± 1.00 |
| Δ from baseline to week four | 4.9 ± 3.5 | 13.9 ± 2.9 | --- | --- |

Furthermore, there was no difference in baseline CRP levels of patients (1.3 ± 1.3 mg/L) and healthy controls (3.8 ± 13.0 mg/L, p = 0.47). Of note, one healthy participant had a CRP level of 97.6 mg/L. Three out of 33 (9%) of patients and 9/58 (16%) of healthy controls had CRP plasma concentrations > 3 mg/L, the cutoff to define low-grade inflammation (Fisher’s exact test’s p = 0.5).

The within-group analysis revealed that in non-responders, CRP plasma levels increased from baseline to the first week of treatment with a medium effect size (d = 0.7, p = 0.01), while in responders, CRP levels at baseline and week one were comparable (Fig.3, Table 2).

**Figure 3.**
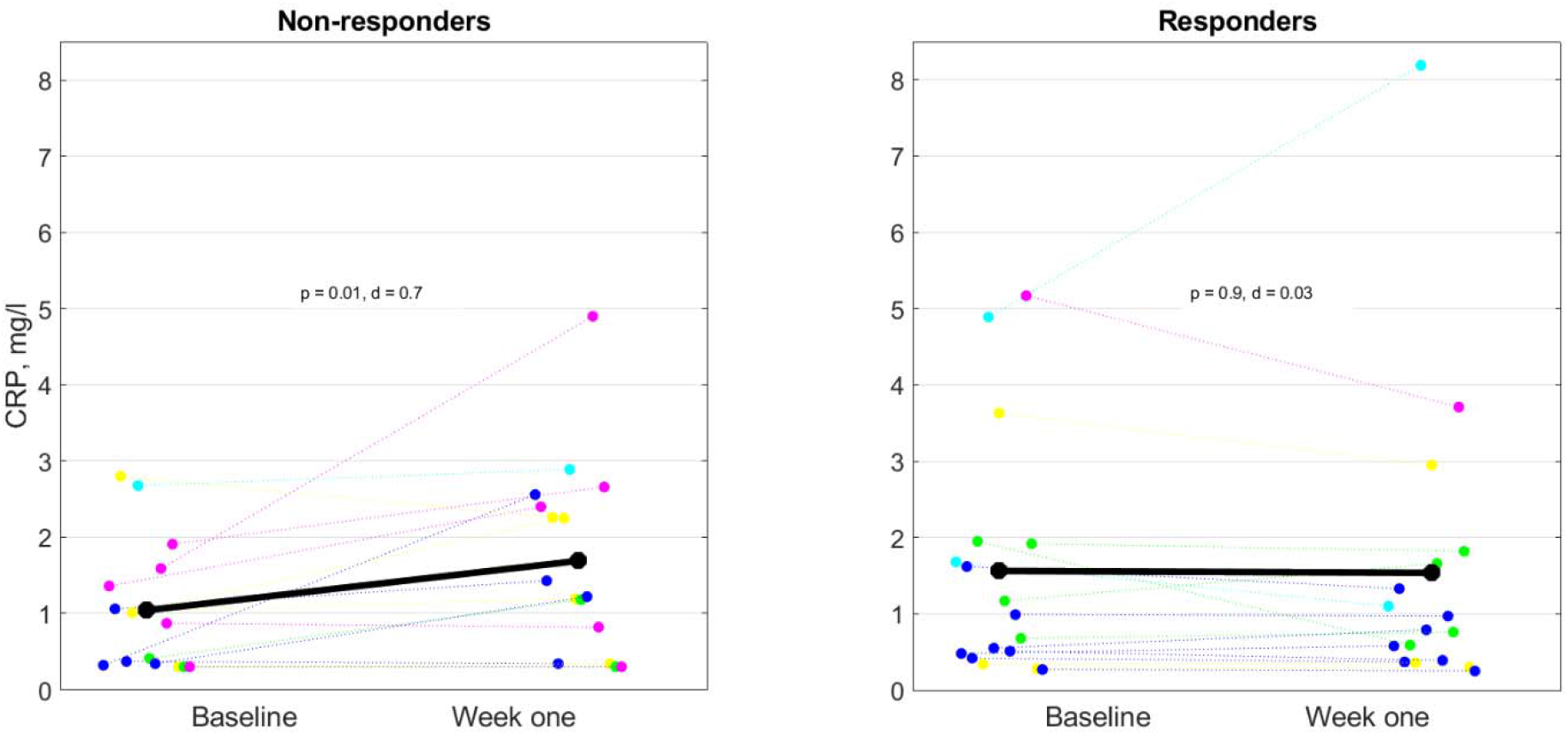
CRP levels at baseline and week one of antidepressant treatment. In non-responders (left), CRP plasma levels increased from baseline to week one of treatment (p = 0.01), while in responders (right), it did not change significantly. The colored circles represent individual patients where different colors correspond to different antidepressant classes as follows: cyan-SNRI, blue-SSRI, yellow-TCA, green-NaSSA. It was not possible to stratify patients by their antidepressant classes due to the small sample size (n = 33). Black circles refer to the average of all patients in a group. The statistical paired t-test was performed using the square root transformed values, while the figure shows non-transformed values to easily see the CRP value of 3 mg/L, the cut-off to define low-grade inflammation.

ΔCRP discriminated between non-responders and responders with the area under the receiver operating characteristic curve of 0.763 ± 0.083 (considered acceptable AUC level, p = 0.010, Fig.4). When cut off at the median of ΔCRP of all patients (ΔCRP = 0.02 mg/L), the test yielded a sensitivity of 69% and a specificity of 70%. The corresponding positive and negative predictive values were 77% and 65%, respectively, and accuracy equalled 70%.

**Figure 4.**
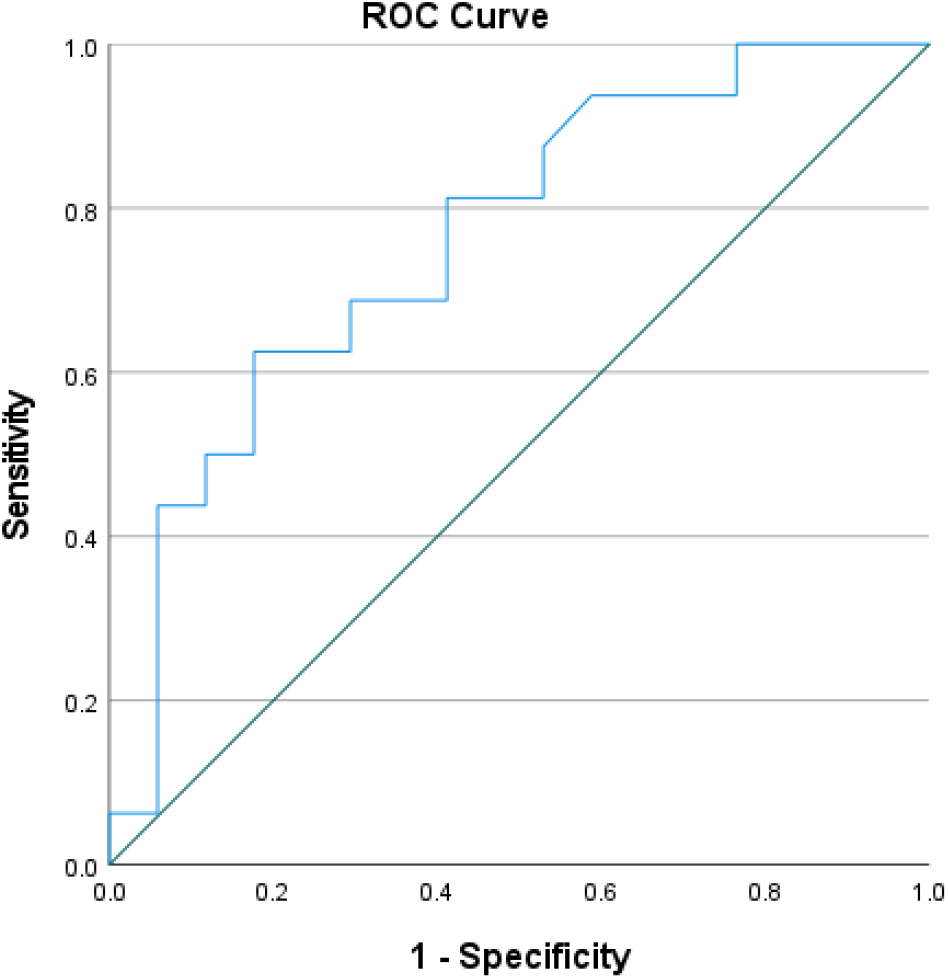
Receiver operating curve. ΔCRP discriminated between the non-responders and responders with the area under the receiver operating characteristic curve of 0.76 ± 0.08 (p = 0.01). A black line represents a discriminative performance at a chance level.

At the individual level, a ΔCRP increase was observed in 13/16 (81%) non-responders and 7/17 (41%) responders (p = 0.03). Even though this result is statistically significant, it is not “predictive” enough to be introduced into clinical setting. Therefore, we performed an additional *post hoc* analysis, which, we believe, is highly motivated clinically. Namely, we combined a ΔCRP increase with an early non-response (defined as a ≤ 25% decrease in the HAM-D score from baseline to week one), where the latter is readily available in clinical practice. We found that a ΔCRP increase together with an early non-response was observed in 13/16 (81%) non-responders and only in 1/17 (7%) responders (p < 0.0001), i.e., this combination provided a strong predictive accuracy relevant both statistically and clinically. For reference, an early ΔHAM-D non-response was observed in 14/16 (87.5%) non-responders and in 7/17 (41%) responders (p = 0.010, Table 3).

**Table 3.** Fisher’s exact test.

| Parameter | Responders | Non-responders | Fisher’s exact test’s p |
| --- | --- | --- | --- |
| ΔCRP increase | 7/17 (41%) | 13/16 (81%) | 0.032 |
| Early ΔHAM-D non-response | 7/17 (41%) | 14/16 (87.5%) | 0.010 |
| ΔCRP increase and early ΔHAM-D non-response | 1/17 (6%) | 13/16 (81%) | <0.0001 |

*A ΔCRP increase was defined as ΔCRP ≥ 0 mg/L from baseline to week one, an early ΔHAM-D non-response was* *defined as ≤ 25% HAM-D decrease from baseline to week one*

## Discussion

This study revealed that MDD patients who do not respond to their antidepressant treatment (i.e., < 50% HAM-D improvement at day 28) present with an increase in the plasma concentration of ΔCRP from baseline to the first week of treatment, while responders do not show significant ΔCRP changes during the early course of treatment. The novelty of the current study is its focus on the within-subject CRP changes (ΔCRP) as a proxy for early antidepressant treatment response, with the future potential for its prediction in clinical settings. The reported effect was observed in 81% of non-responders and 41% of responders, which limits its clinical usefulness. Yet, when ΔCRP was used in the combination with early (7-day) ΔHAM-D, a ΔCRP increase and no symptom improvement at day 7 was observed in 81% of non-responders and only 6% of responders, which is both statistically significant and clinically relevant.

Common non-response rates of about 30 – 50% (in the current study – 16/33 (48%)) for any given antidepressant class suggest that the monoamine hypothesis of MDD (that explains MDD by the depletion of monoamines, e.g., serotonin) alone is insufficient to explain the MDD pathology (Pastis et al., 2024). The immunological model of MDD tries to explain some of its symptoms (e.g., anhedonia, a lack of motivation, fatigue, psychomotor slowing, reduced food intake) as the non-specific reaction of the organism to infection and inflammation (Young et al., 2016). It suggests that in a subset of patients with a unique symptom presentation and treatment response, the disease is primarily driven by inflammation. Interestingly, the interventions that are known to alleviate depression (e.g., physical exercise, mindfulness or psychotherapy) also reduce peripheral inflammatory biomarkers (Rantala et al., 2018).

According to meta-analyses, 25 – 33% of MDD patients exhibit low-grade inflammation as defined by the standard cutoff of the inflammation presence, i.e., the CRP levels > 3 mg/L (Miller et al., 2025). Notably, the large case-control study counting 26,894 MDD patients and 59,001 controls reported CRP > 3mg/L in 21% of patients but also in as many as 17% of the controls (Pitharouli et al., 2021). In the present study, 9% of patients and 16% of healthy controls had CRP plasma concentrations > 3 mg/L. This evidence suggests that cross-sectional CRP levels at baseline can not be used for MDD diagnosis or prediction of antidepressant treatment response. This suggestion is also supported by the longitudinal study that included 340,882 Swedish individuals, reporting that CRP ≥ 4mg/L at baseline did not predict later MDD occurrence (Zeng et al., 2024).

Regarding antidepressant treatment, some studies observed elevated levels of CRP and related pro-inflammatory cytokines over the whole course of treatment of several weeks in MDD patients (Strawbridge et al., 2018; Köhler et al., 2018). Others report that antidepressants decrease several markers of peripheral inflammation. The meta-analysis found no association between reductions in peripheral inflammation and antidepressant treatment response although few studies reported it (Köhler et al., 2018). In view of heterogeneous findings on the ability of the baseline CRP and other inflammation markers to predict treatment response (Young et al., 2016; Strawbridge et al., 2018), it has been suggested that low-grade inflammation plays a role in the poor responsiveness to some but not other classes of antidepressants. For example, patients with SSRI-resistant depression have higher levels of interleukin-6 and tumor necrosis factor-α compared to normal controls, and formerly SSRI-resistant patients (O’Brien et al., 2007). At the same time, antidepressants with noradrenergic, dopaminergic, or glutamatergic action reduce inflammation by restoring monoaminergic signaling and neurotrophin function and reducing central cytokine signaling, microglial activation and activity of the stress axes (Young et al., 2016; Strawbridge et al., 2018). Due to the small sample size, here, we could not control our analysis for the effect of the antidepressant class. Nevertheless, the fact that we observed an early ΔCRP effect despite the heterogeneity of the used antidepressants suggests that early effects of antidepressants on ΔCRP might be less relevant. Possible reasons for that include unspecificity of early antidepressant treatment, such as adverse drug effects (e.g., SSRI-induced nausea), or its beneficial effect on sleep (e.g., low doses of some antidepressants improve insomnia, while they do not have an antidepressant effect at that time (Rosenblum et al., 2025)). At the same time, we should stress that the CRP levels of our patients were below the threshold (< 3 mg/L, Fig.3 and Table 2); thus, we also did not expect to observe large differences. Likewise, we can not exclude possible therapeutic effects of the in-patient setting (e.g., stress reduction). Importantly, our study was not designed to assess the antidepressant-specific effects of ΔCRP; its aim was to assess whether early change in CRP levels predict HAM-D changes by whatsoever. Future studies should measure late changes in CRP in patients stratified by their antidepressant classes.

Notably, while ΔCRP differed between responders and non-responders, the baseline CRP values as well as CRP concentrations after one week of treatment were comparable in both subgroups. This could be explained by the influence of comorbidities, such as peripheral inflammatory processes, acute or subclinical infections, chronic diseases, allergy and lifestyle covariates, such as diet, smoking and physical activity (Young et al., 2016), which are difficult or sometimes even impossible to control for. Here, the analysis performed without an adjustment for covariates showed that higher CRP levels at week one of treatment corresponded to higher depression severity. The effect disappeared after the correction for age, gender and BMI, which is in line with the larger study in 112 patients with depression that found a positive association between CRP levels and symptom severity, which disappeared after controlling for BMI (Krogh et al., 2014). We believe that the repeated measures within-subject analyses used in our study have controlled for comorbidities and inherent heterogeneity of MDD. In view of our findings, we propose that future research should focus on the within-subject ΔCRP instead of comparing between-group absolute CRP values as has been done before. In other words, our findings suggest that the CRP reactivity is more important than its baseline levels.

Regarding the possible mechanisms, there is evidence that some proinflammatory cytokines (e.g., interleukin 1α and 1β) that normally stimulate the hypothalamic-pituitary-adrenal (HPA) axis, might also promote glucocorticoid resistance within the immune system (Young et al., 2014). Normally, cortisol, a glucocorticoid released by the HPA axis and the main stress hormone, suppresses the inflammatory response, e.g., by inhibiting further cytokine production. In chronic depression, glucocorticoid resistance may disrupt the negative feedback loop to the HPA axis and the immune system, resulting in long-term inflammatory states and hyperactivity of the HPA axis, which in turn maintain depressive symptoms, including mood changes, cognitive impairments, and sleep disturbances (Kim et al., 2022).

Another evidence suggests that proinflammatory cytokines activate the enzyme indoleamine-2,3-dioxygenase (Young et al., 2016). This activation leads to a decrease in serotonin production and an increase in the production of kynurenic and quinolinic acids (Wium-Andersen et al., 2013). The latter results in increased glutamate release and thereby decreased production of trophic factors, including the brain-derived neurotrophic factor (BDNF). Suppressed BDNF secretion may result in impaired neuroplasticity and neurogenesis. Accordingly, inflammation may contribute to depression via the suppression of BDNF and poor neuroplasticity, as it has been formulated in the “neurotrophic hypothesis of depression” (Madsen et al., 2024).

Most studies agree that brain BDNF has an essential role for the efficacy of antidepressant treatments (Madsen et al., 2024). Interestingly, an early reduction (or non-rise) in CRP mirrors an early rise in BDNF. For example, an early increase in plasma BDNF levels after treatment initiation predicted antidepressant response, while the lack thereof predicted non-response (Dreimüller et al., 2012). Furthermore, an early increase in BDNF serum levels translated to an improvement in cognitive abilities by antidepressant treatment (Mikoteit et al., 2015). It should be noted, however, that even though there is evidence that both CRP and BDNF reflect antidepressant treatment response, currently, their link is hypothetical and future studies that measure CRP and BDNF simultaneously are needed to further understand their relationships and more generally, the inflammatory and neurotrophic hypotheses of depression.

This study has several limitations. First, it has a relatively small sample size of the patients treated with various antidepressants with possibly differential effects on inflammation, meaning that our findings should be replicated by future larger studies. However, as we do not refer to final CRP levels, early effects of antidepressants on CRP might be less relevant, as week one is too early for most antidepressants to evaluate their final impacts. Second, all analyses were correlational, precluding us from a discussion on causal relations between inflammation (as reflected by CRP levels) and antidepressant response and leaving the possibility that this association is due to an unknown confounding factor. Third, even though we adjusted the between-subject ΔCRP analysis for several covariates known in the literature, we cannot exclude the possibility of residual confounding (e.g., acute infections at the time of blood measurements, chronic diseases, lifestyle covariates, including smoking and covariates related to socioeconomic status). Nevertheless, given that besides the between-subject analysis we also performed the repeated measures within-subject analysis expected to control for such residual confounding, our results are unlikely to be chance findings.

In summary, our study revealed that non-response to antidepressant treatment is associated with an increase in the plasma ΔCRP level from baseline to the first week of treatment. When we used ΔCRP as the only early marker of response, an increase in ΔCRP was observed in 81% of non-responders and 41% of responders, which seems not sufficient for the introduction of ΔCRP to clinical settings. When we used ΔCRP in combination with 7-day ΔHAM-D, we found that a ΔCRP increase together with an early non-response was observed in 81% of non-responders and only 6% of responders, which is much more relevant clinically. Namely, the use of the combined marker (ΔCRP and early clinical response) might allow clinicians to identify patients at risk of non-response and consider intensifying treatment (e.g., by increasing the dose, switching medication, or initiating augmentation strategies) earlier than usual (i.e., without waiting until weeks 4–6 as is the common practice now).

Furthermore, given that different markers are interrelated in a complex fashion, we expect that if ΔCRP is used in combination with other predictive biomarkers (e.g., BDNF changes, HRV and cordance during sleep), in the future, they a) could be introduced into daily psychiatric practice to modify treatment plans according to personalized histories and peripheral biomarker results; and b) could be used to identify the inflammatory subtype of MDD and pave the way for personalized treatments that target the inflammatory pathways, potentially leading to more effective interventions.

## Data Availability

All data produced in the present work are contained in the manuscript

## Acknowledgments

We would like to thank Prof. Dr. Thomas Pollmächer for reviewing this Manuscript and for his constructive input. YR and MD were supported by the Dutch Research Council (NWO).

